# Screening for SARS-CoV-2 persistence in Long COVID patients using sniffer dogs and scents from axillary sweats samples

**DOI:** 10.1101/2022.01.11.21268036

**Authors:** Dominique Grandjean, Dorsaf Slama, Capucine Gallet, Clothilde Julien, Emilie Seyrat, Marc Blondot, Judith Elbaz, Maissa Benazaiez, Dominique Salmon

## Abstract

**Objectives:** Dogs can be trained to identify several substances not detected by humans, corresponding to specific volatile organic compounds (VOCs). The presence of VOCs, triggered by SARS-CoV-2 infection, was tested in sweat from Long COVID patients.

**Patients and methods:** An axillary sweat sample of Long COVID patients and of COVID-19 negative, asymptomatic individuals was taken at home to avoid any hospital contact. Swabs were randomly placed in olfaction detection cones, and the material sniffed by at least 2 trained dogs.

**Results:** Forty-five Long COVID patients, mean age 45 (6-71), 73.3% female, with prolonged symptoms evolving for a mean of 15.2 months (5-22) were tested. Dogs discriminated in a positive way 23/45 (51.1%). Long COVID patients versus 0/188 (0%) control individuals (p<.0001).

**Conclusion:** This study suggests the persistence of a viral infection in some Long COVID patients and the possibility of providing a simple, highly sensitive, non-invasive test to detect viral presence, during acute and extended phases of COVID-19.

## Introduction

Dogs have olfactory capacities several hundred times greater than humans, likely due to a much larger zone of olfactory epithelium with 40 times more olfactory cells, and to the presence of the Jacobson vomero-nasal organ, which does not exist in humans. Thanks to their intelligence, sociability, and high learning ability, dogs can be trained to detect a comprehensive number of different substances both from biological (plant, animal, or human odors) or non-biological origin (chemical or technological products).

Since April 2020, the veterinarians of the National Veterinary School of Alfort (ENVA) have been training dogs to detect Sars-CoV-2 virus in human sweat, by detecting volatile organic compounds (VOCs) in infected patients [1]. The VOCs exact nature is still under identification [2]. During the acute COVID-19 phase, the first results show a detection sensitivity close to 95% and a specificity of 97% for confirmed cases (positive PCR) versus asymptomatic and negative PCR subjects [3,4].

For Long COVID patients, the persistence of RNA and/or viral proteins is a widely discussed hypothesis. The literature has documented the viral RNA persistence in olfactory slots [5], digestive tissue sections [6], brain [7] and viral proteins persistence in monocytes [8].

Therefore, it is of great scientific interest to assess whether dogs can identify SARS-CoV-2 persistence in Long COVID patients, as they do in the initial phase of the disease.

## Method

The included patients had all a symptomatic initial episode of COVID-19, then prolonged symptoms which corresponding to the 2021 WHO definition of post-acute covid syndrome [9]. They were followed at Hotel Dieu Hospital in a descriptive cohort for which an Ethics approval was granted (Institutional Review Board of Mondor, Creteil, France, IRB 00011558, Approval number 2020-088). Patients were asked to take an axillary sweat sample at home to avoid any hospital contact and potential parasite odors. Before any shower in the morning, the patients had to apply a sterile surgical swab under each armpit for 5 minutes, slip it into a freezer bag and send it after sealing it by mail. At ENVA, swabs were placed in a glass container in an olfaction detection cone according to the Nosaïs procedure (Figure 1), [1,3]. Each case to be checked was randomly placed with 4 negative swabs from COVID-19 negative and asymptomatic individuals in 5 different cones. The material was sniffed by 2 different dogs trained and validated to sniff and discriminate positive samples for SARS-CoV- 2. The dog and his handler were both blinded to the Long COVID sample location.

**Figure 1:**
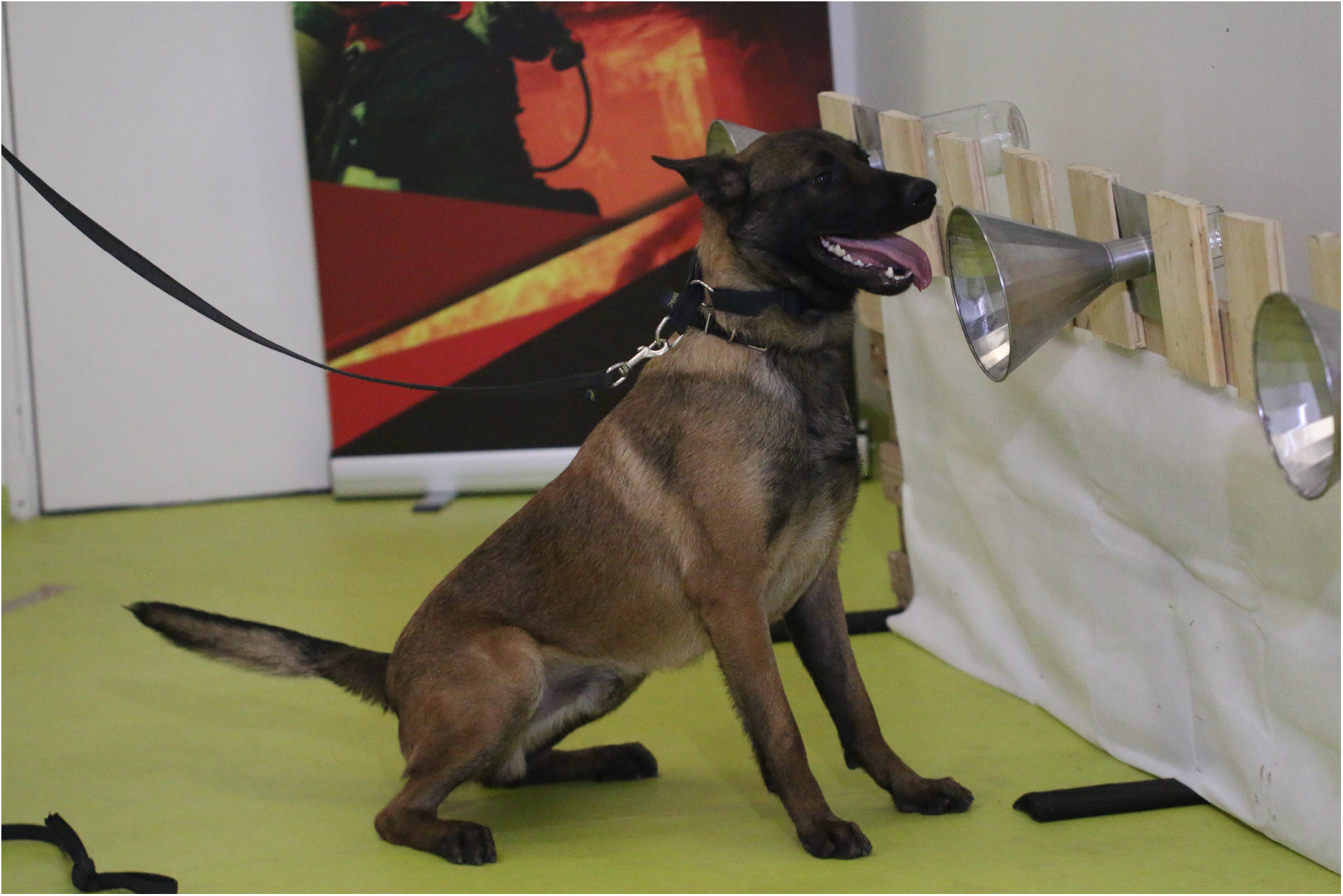
Dog sniffing setting: A handler is responsible of for the guidance of the dog. The sniffing cone helps the dog sniff the scent of the samples put inside glass containers, which are held in empty boxes.

## Results

### Characteristics of the population

Between May and October 2021, 45 Long COVID patients sent their samples to ENVA. Average age was 45 (6-71) and 73.3% were female. No patient had been admitted in intensive care unit during the acute phase.

Prolonged symptoms had been evolving for an average of 15.2 months (range: 5-22). Main symptoms of prolonged phase were intense fatigue (n=37, 82.2%), neurocognitive disorders such as concentration and attention difficulties, immediate memory loss (n=24, 53.3%), myalgias/arthralgias (n=22, 48.9%), cardiopulmonary symptoms (dyspnea, cough, chest pain, palpitations) (n=21, 46.7%), digestive symptoms (diarrhea, abdominal pain, reflux, gastroparesis…) (n=18, 40.0%), ENT disorders (hyposmia, parosmia, tinnitus, nasal obstruction, inflammatory tongue, dysphonia, sinusitis) (n=18, 40.0%) (table 1). 11 (24.4) patients had at least one positive SARS-CoV-2 serology before any vaccination, 29 (64.4%) had a negative SARS-CoV-2 serology and 5 (11.1%) had no serology results.

**Table 1:**
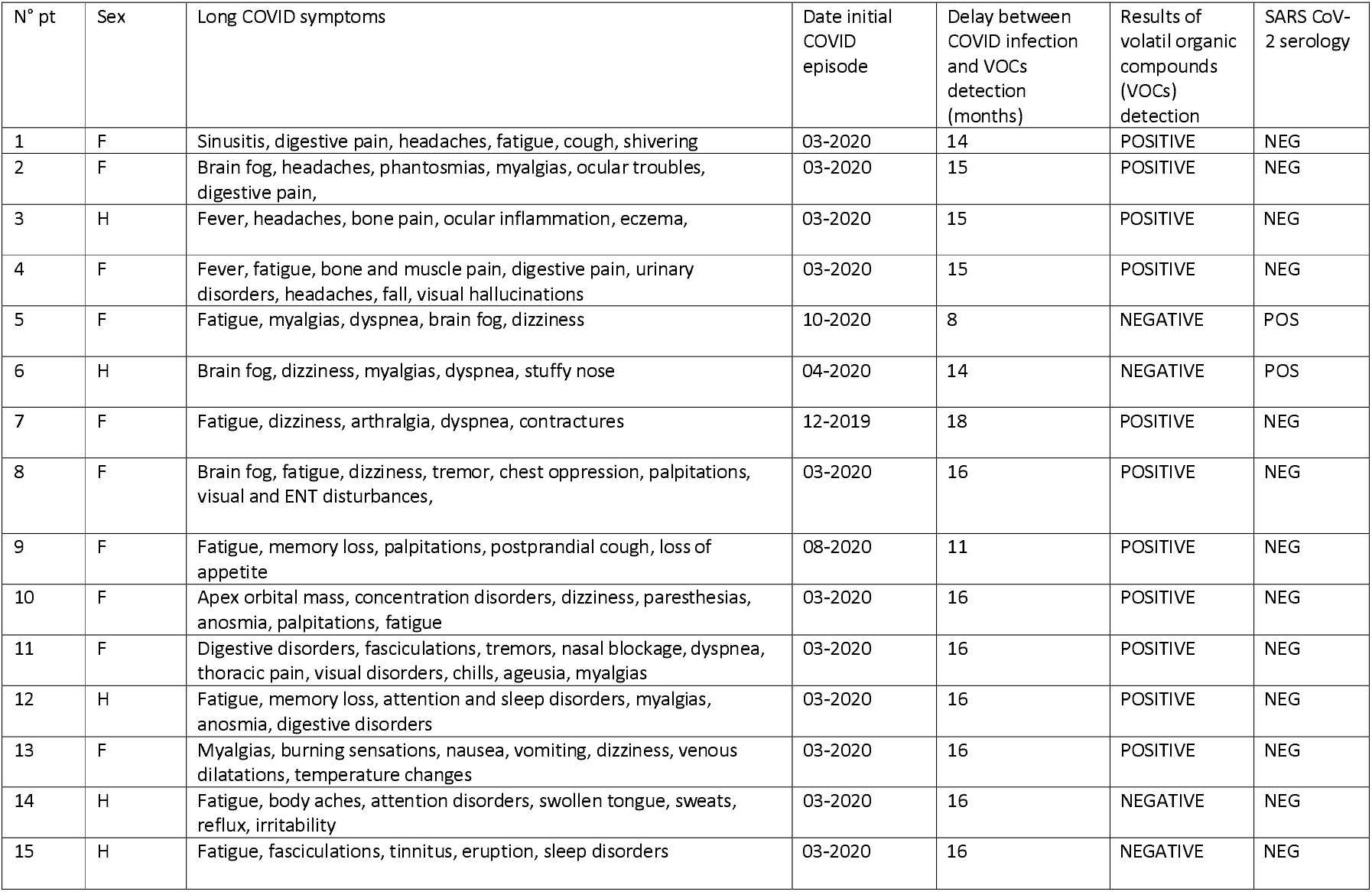
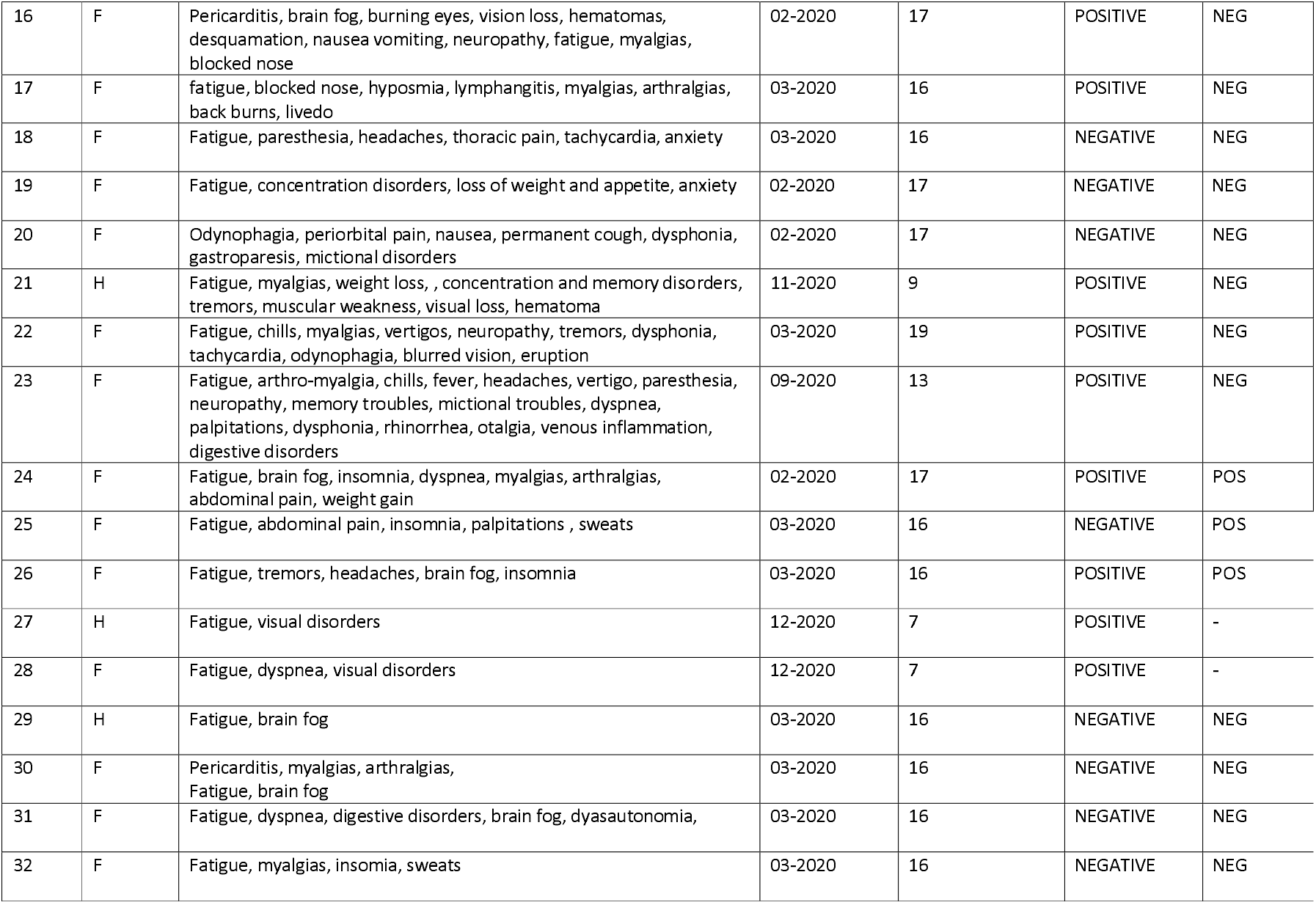
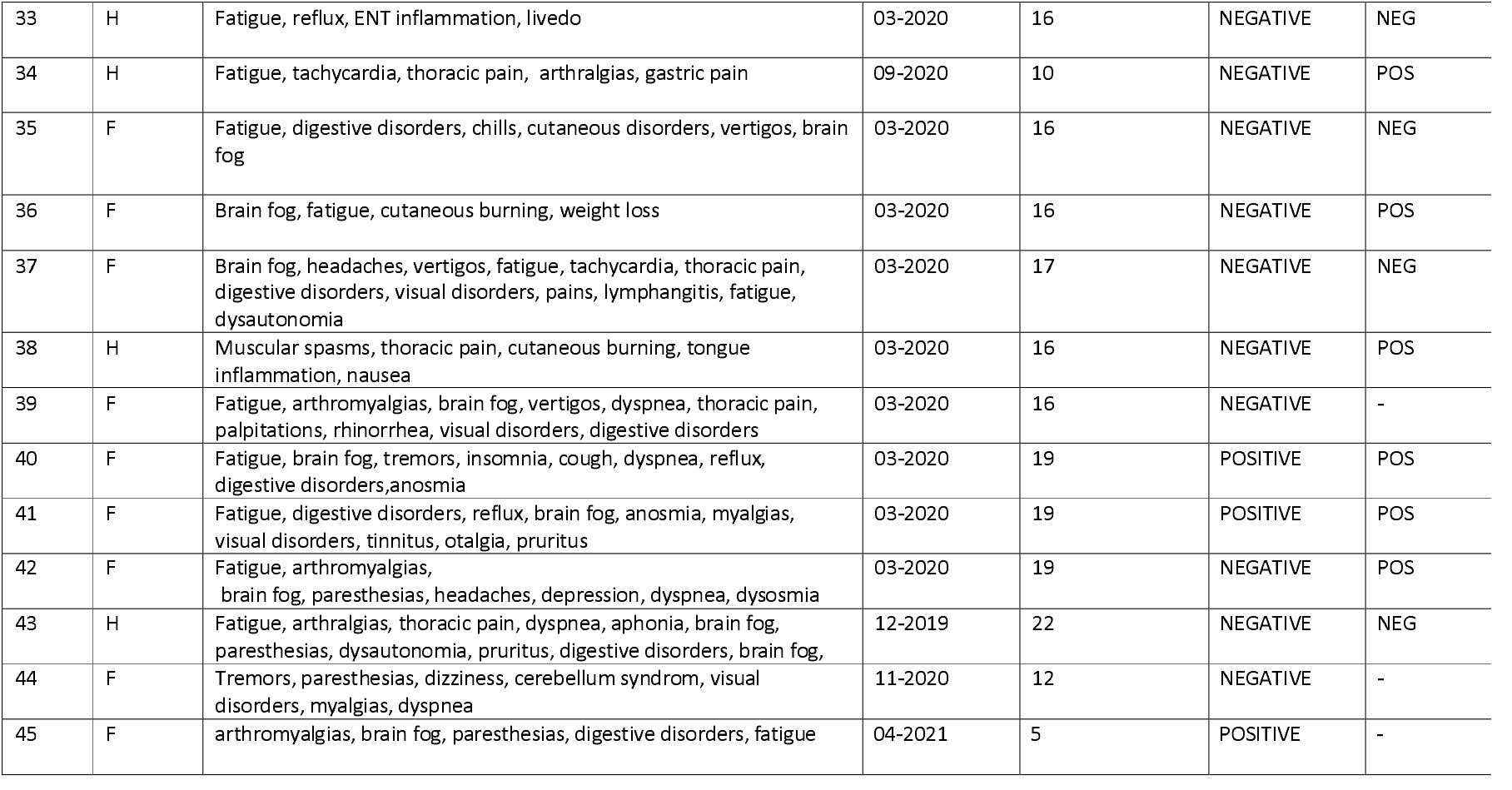
Characteristics of the Long COVID patients included in the study and result of volatile organic compounds (VOCs) detection

### Dog discrimination

23 long COVID samples out of 45 (51.1%), were discriminated in a positive way while none out of 188 (0%), was discriminated among control samples (p<.0001). There was no significant difference in the rate of positive discrimination between subjects with a positive SARS-CoV-2 serology (4/11, 36.4%) and those with a negative serology (16/29, 55.2%), (p=0.478).

## Discussion

This study, performed in Long COVID patients, shows for the first time that dogs can detect volatile organic compounds (VOCs) up to 1.5 year after the initial phase of COVID-19. These results strongly suggests that SARS-CoV-2 can persist at least in some Long COVID patients.

During the acute initial phase of COVID-19, the performance of canine SARS-CoV-2 detection was shown to be excellent. A 1^st^ study was performed in Recife, Brazil in which 100 hundred volunteers with COVID-like symptoms participated, and both axillary sweat samples for dog detection and nasopharynx/oropharynx swabs for qPCR were collected. Two dogs, previously trained, detected 97.4% of the sweat samples positive with COVID-19 PCR, including a false-negative qPCR-test; the positive predictive value was 100% and the negative predictive value was 98.2% [3]. A 2^nd^ study was conducted in Lebanon in 256 patients with a proven COVID-19 (PCR positive) and in 203 PCR negative and asymptomatic subjects. Sweat samples were obtained within the 24 hours of the PCR testing and independently tested by two dogs. This detection by dogs showed a sensitivity of 100% and a specificity of 98.6% [4].

In this study with Long COVID patients, it is difficult to interpret negative discriminations as the sample quality take at home may have varied, and postal delays may have altered the preservation. Therefore, the sensitivity of canine olfactory test might be underestimated. Nevertheless, in this study, the specificity of SARS-CoV-2 olfactory detection by dogs is strikingly high, like in previous studies [3, 4].

Interestingly, the finding that trained dogs can discriminate sweats in Long COVID patients corroborates the previous cases of persistence of SARS-CoV-2 RNA and/or antigens, documented in the literature for long Covid patients [5-8]. Up to date, it had not been demonstrated whether it corresponds to the replicative virus or not. This canine detection test, with detection of volatile organic compounds, supports the hypothesis that the virus is still actively replicating. The nature of these VOCs is currently being identified by several international laboratories in different countries. The training of the dogs now includes the uses of lures - produced by Pasteur Institute (Paris, France)- that are under validation by several international teams (ongoing publications). These lures are derived from supernatants of inactivated SARS-CoV-2 viral cultures, treated with trypsin, containing both viral proteins and volatile compounds, that dogs learn to discriminate.

It has been recently shown that trained dogs can also distinguish SARS-CoV-2-infections from other viral infections with mean specificities above 90% [10].

The strengths of this study are the presence of negative controls during each test, the fact that all controls were all negatively discriminated and the easy VOCs detection method. This very sensitive and non-invasive technique, which can be used both during acute and extended phases, is also faster than the RT-PCR and provides a holistic information. Indeed, it gives reliable results on the existence of a persistent infection, even when the precise location of the virus reservoir in the body is not known.

We also recognize some limitations due to the relatively small sample size of our population, and the possible heterogeneity of self-sampling done by patients at home that can lead to the existence of false negatives canine olfactory detection (which minimizes test sensitivity).

In conclusion, these results confirm the high probability of a SARS-CoV-2 viral persistence at least for some Long COVID patients, possibly with virus actively replicating. It also shows the limited utility of serological tests made during Long COVID. Such conclusions are of major importance for the management of future Long COVID treatments trials. In addition, with a better characterization of the detected VOCs, an improvement of odor sampling methods and the development of point-of-care instruments, this detection by dogs could also help to implement scent-based tests for other major human pathogens.

## Data Availability

All data produced in the present study are available upon reasonable request to the authors

## References

1 Grandjean D, Sarkis R, Lecoq-Julien C, Benard A, Roger V, Levesque E, Bernes-Luciani E, Maestracci B, Morvan P, Gully E, Berceau-Falancourt D, Haufstater P, Herin G, Cabrera J, Muzzin Q, Gallet C, Bacqué H, Broc JM, Thomas L, Lichaa A, Moujaes G, Saliba M, Kuhn A, Galey M, Berthail B, Lapeyre L, Capelli A, Renault S, Bachir K, Kovinger A, Comas E, Stainmesse A, Etienne E, Voeltzel S, Mansouri S, Berceau-Falancourt M, Dami A, Charlet L, Ruau E, Issa M, Grenet C, Billy C, Tourtier JP, Desquilbet L. Can the detection dog alert on COVID-19 positive persons by sniffing axillary sweat samples? A proof-of-concept study. PLoS One. 2020 Dec 10;15(12):e0243122

2 Cambau E, Poljak M. Sniffing animals as a diagnostic tool in infectious diseases. Clin Microbiol Infect. 2020 Apr;26(4):431–435.

3 Maia RCC, Alves LC, da Silva JES, Czyba FR, Pereira JA, Soistier V, Julien CL, Grandjean D, Soares AF. Canine Olfactory Detection of SARS-COV2-Infected Patients: A One Health Approach. Front Public Health. 2021 Oct 21;9:647903. doi: 10.3389/fpubh.2021.647903.

4 Sarkis R, Lichaa A, Mjaess G, Saliba M, Selman C, Julien C, Grandjean D, Nabil M. Jabbour N.M. New method of screening for COVID-19 disease using sniffer dogs and scents from axillary sweat samples. Journal of Public Health, 2021, Jun 23;fdab215. doi: 10.1093

5 De Melo GD, Lazarini F, Levallois S, Hautefort C, Michel V, Larrous F, Verillaud B, Aparicio C, Wagner S, Gheusi G, Kergoat L, Kornobis E, Donati F, Cokelaer T, Hervochon R, Madec Y, Roze E, Salmon D, Bourhy H, Lecuit M, Lledo PM. COVID-19-related anosmia is associated with viral persistence and inflammation in human olfactory epithelium and brain infection in hamsters. Sci Transl Med. 2021 Jun 2;13(596):8396

6 Cheung CCL, Goh D, Lim X, Tien TZ, Lim JCT, Lee JN, Tan B, Tay ZEA, Wan WY, Chen EX, Nerurkar SN, Loong S, Cheow PC, Chan CY, Koh YX, Tan TT, Kalimuddin S, Tai WMD, Ng JL, Low JG, Yeong J, Lim KH. Residual SARS-CoV-2 viral antigens detected in GI and hepatic tissues from five recovered patients with COVID-19. Gut. 2022 Jan;71(1):226–229. doi: 10.1136/gutjnl-2021-324280.

7 Chertow D, Stein S, Ramelli S, Chung J-Y, Singh M, Yinda K, Winkler C, et al. SARS-CoV-2 infection and persistence throughout the human body and brain. Dec 2021. https://www.researchsquare.com/article/rs-1139035/v1

8 Patterson BK, Guevara-Coto J, Yogendra R, Francisco EB, Long E, Pise A, Rodrigues H, Parikh P, Mora J, Mora-Rodríguez RA. Immune-Based Prediction of COVID-19 Severity and Chronicity Decoded Using Machine Learning. Front Immunol. 2021 Jun 28;12:700782. doi: 10.3389/fimmu.2021.700782.

9 WHO definition of post acute covid syndrome. WHO/2019-nCoV/Post_COVID-19_condition/Clinical_case_definition/2021.1. Dernière consultation le 27/12/2021

10 Ten Hagen NA, Twele F, Meller S, Jendrny P, Schulz C, von Köckritz-Blickwede, M, Osterhaus A, Ebbers H, Pink I, Welte T, Manns MP, Illig T, Fathi A, Addo MM, Nitsche A, Puyskens A, Michel J, Krause E, Ehmann R, von Brunn A, Ernst C, Zwirglmaier K, Wölfel R, Nau A, Philipp E, Engels M, Schalke E, Volk HA. Discrimination of SARS-CoV-2 Infections From Other Viral Respiratory Infectionsby Scent Detection Dogs. Front Med (Lausanne). 2021 Nov 18;8:749588. doi:10.3389/fmed.2021.749588

